# Quadrangular Space Syndrome: A Systematic Review of Surgical and Medical Therapeutic advances

**DOI:** 10.1101/2022.05.21.22275366

**Authors:** Sundip Charmode, Shelja Sharma, Sudhir Kushwaha, Simmi Mehra, Shalom Philip, Ranjna Janagal, Pratik Amrutiya

## Abstract

The Axillary Nerve (AXN) and Posterior Circumflex Humeral Artery (PCHA) are compressed in quadrangular space syndrome (QSS), which can be treated with conservative approaches or surgical decompression in recalcitrant instances. There are no clear guidelines for determining which surgical method is optimal for treating QSS and other disorders that mirror QSS. The goal of this study is to grade and review past, current, and planned medicinal and surgical care modalities for QSS. PROSPERO was used to submit the review protocol (ID: 332766). To identify recent advances in the methods/techniques of medical and surgical management of QSS, PubMed and Medline databases were searched until March 2022 for publications, including case studies, case reports, and review articles, using Medical Subject Headings (MeSH) terms like quadrilateral space syndrome, surgical management, and medical management. Throughout the study, all the authors scrupulously followed a well-developed registered review process and the risk of bias in systematic reviews (ROBIS) guidance tool. Data on proposed medical and surgical management methods/techniques was compiled, and each was analyzed based on the underlying neuro-vascular systems. There were 88 items found in the first search. Following applying the inclusion and exclusion criteria, 16 papers were chosen for synthesis in the review study after a thorough assessment. Three studies (conservative and advanced) focused on medical care of QSS, while 12 articles (prior, current, and newer) focused on surgical management of QSS. Only four of the 15 studies reviewed proposed different surgical approaches/techniques for surgical decompression in QSS. There were two regularly used surgical procedures discovered, one anterior/delto-pectoral and the other posterior/scapular. The anterior route is more technically straightforward and can be employed for surgical QSS decompression.

## 1. Introduction

Quadrilateral Space Syndrome (QSS) is a rare neuro-vascular entrapment disease in which the Axillary nerve (AXN) or Posterior Humeral Circumflex Artery (PHCA) are entrapped in the quadrilateral space due to injury, fibrous bands, or muscle border hypertrophy [1].

The disease typically affects young adults aged 20 to 35, particularly athletes who participate in overhead sports such as volleyball [2], baseball [3], swimming [4], and other activities that require regular abduction and external rotation, such as yoga [5] or window washing [6]. The subscapularis muscle and shoulder capsule border the intermuscular space on the superior side, and the teres major muscle on the inferior side. The long head of the triceps and the surgical neck of the humerus limit it medially and laterally [7]. Loose connective tissue, fat, veins, the AXN, and the PHCA are all included [7]. Patients are usually advised to start with conservative therapies like physical therapy and activity modification [2]. When patients do not respond to conservative therapy for at least six months, surgical decompression is considered [8]. The management entails decompression, which entails a variety of techniques that have yet to be validated, particularly among the Indian population. The goal of our research is to grade the various medicinal and surgical treatments for Quadrangular Space Syndrome.

### Functional Anatomy of Axillary Nerve (AXN)

The AXN is located behind the brachial artery and lateral to the radial nerve, in front of the subscapularis. The AXN joins the posterior circumflex humeral artery in the quadrangular space, positioned between the lateral and long heads of the triceps muscle on the inferior side of the subscapularis. The anterior branch of AXN innervates the deltoid muscle, while the posterior branch innervates the teres minor and deltoid. The posterior branch also innervates the skin across the inferior two-thirds of the deltoid muscle’s posterior surface [9].

### Functional Anatomy of Posterior Humeral Circumflex Artery (PHCA)

It enters the posterior scapular region after passing via the quadrangular space. It separates into anterior and posterior branches within the quadrangular space, looping antecedently around the humeral surgical neck to give blood to the superior, inferior, and side sections of the humeral head, the glenohumeral joint, and the surrounding shoulder muscles [10,11]. The borders of the quadrangular space, as well as the structures that cross through it, are depicted in Figure 1.

**Figure 1.**
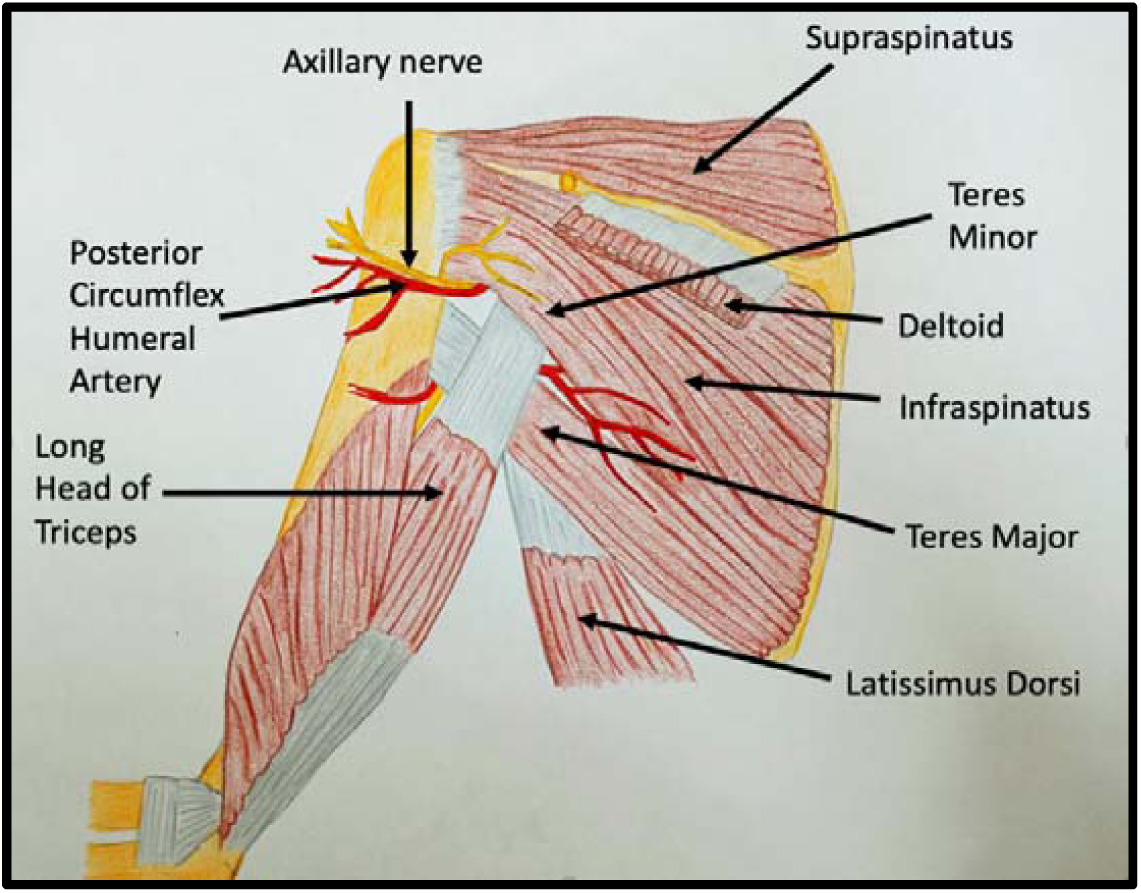
Boundaries of Quadrangular space with its contents. (This image has been created and edited by Dr. Shalom Philip, Senior resident, AIIMS Rajkot)

## 2. Materials and Methods

### REVIEW

#### METHODS

To reduce the risk of bias in the study, a systematic review protocol was prepared and submitted with PROSPERO at the Centre for Reviews and Dissemination, University of York (ID: 332766). The review protocol can be accessed from https://www.crd.york.ac.uk/prospero/.

### 2.1 Eligibility criteria

The articles which satisfied the inclusion and exclusion criteria were eligible for review. The inclusion criteria were articles written by both Indian and foreign authors until 31st March 2022, articles related to all relevant medical methods and surgical methods of management of QSS, articles experimenting new methods of treatment on animals for human applications were included. Excluded articles included those which were published after 31st March 2022 and the articles focusing on topics other than medical and surgical methods of management of QSS. There was restriction for non English language of publication. The publications were divided into two groups/Table 2 and 3, which represented articles focusing on medical and surgical management techniques, respectively.

### 2.2 Search Strategy

A scoping review was conducted of articles published till March 2022 on PubMed, and Medline using these MeSH terms: Quadrilateral Space Syndrome; Quadrangular Space Syndrome; Surgical management; Medical management; Neurovascular structures; Throwing Athletes. The citation search was carried out for all the selected articles in study.

### 2.3 Study Selection

After applying inclusion and exclusion criteria, two authors (SP and SC) independently assessed all of the titles, abstracts, full text articles, and reviews found during the initial search, and relevant publications were shortlisted. All the shortlisted full publications were downloaded and independently examined for relevant data using the data extraction checklist prepared by both authors. The authors and the methods of the investigations were not hidden from the reviewers. Any differences were settled through conversation or the involvement of a third reviewer (SM). The special data needed for the review is mentioned on the checklist (Table 1). We chose and incorporated the articles that contained this information.

**Table 1.** Checklist for Data Extraction from the Selected Articles.

| Sr. no | Data Collected | Data Ignored |
| --- | --- | --- |
| 1 | Details of various older and recent methods (conservative as well as advanced) of medical management of QSS | Variations in the arrangement/course of neurovascular structures (AXN and PHCA) passing through the Quadrangular Space |
| 2 | Different procedures/methods used for surgical management of QSS like surgical decompression, etc. | Data regarding the variations in the clinical presentation of patients having QSS |
| 3 | Details regarding the various older and newer techniques of surgical decompression used to treat QSS. | Data regarding various other conditions mimicking the signs and symptoms of QSS |
| 4 | Disadvantages of older methods/techniques used for medical and surgical decompression of QSS |  |
| 5 | Advantages of newer methods/techniques used for medical and surgical decompression of QSS |  |

QS: Quadrangular space; QSS: Quadrangular space syndrome

### 2.4 Data Collection process

The authors created a data extraction form that they used to collect data from any two papers they choose, and it was verified after the pilot trial. Two reviewers (SP and SC) worked separately to gather data from all the studies that were included. Disagreements were addressed through dialogue and the participation of a third independent reviewer (SM). The following features of the study were gathered: (i) the research author; (ii) the study design; (iii) the country and year of publication; (iv) the number of participants; (v) the participants’ age group; (vi) the participants’ gender; and (vii) the participants’ ethnicity. The following information was gathered about the methods/techniques used for the medical and surgical management of QSS: (i) the previous, existing, and recent methods used for medical and surgical management of QSS; (ii) the limitations of the older and existing methods/techniques used for medical and surgical management of QSS (iii) the advantages of the various recent and upcoming methods/techniques used for medical and surgical management of QSS. Checklist (Table 1).

### 2.5 Risk of Bias Assessment

Two reviewers (SP and SC) independently conducted the risk of bias assessment that was included in the Data Extraction Form. The risk of bias in systematic reviews (ROBIS) tool was used to assess the risk of bias in our review study [12].

## 3. Results

### 3.1 Literature search

There were 88 publications found after the initial search in PubMed database. After filtering for the English language, original content, and human involvement, 66 remained. Duplicate articles (n= 2) were deleted, and two reviewers (SP and SC) independently assessed these 66 publications. The same two independent reviewers independently examined 8 papers found through citation searches for eligibility against the pre-specified inclusion criteria. Disagreements were settled by conversation. After applying the inclusion and exclusion criteria, 16 articles remained. 38 publications were excluded due to irrelevant text and 12 publications due to unavailability of full text. Based on each author’s appraisal and cross-verification, 15 publications were selected for synthesis. One review article namely Nicholas Dalagiannis, et al., 2020 [13] was eligible for review based on the inclusion and exclusion criteria but was eliminated after reviewers screened them for irrelevant data that was outside the scope of the current review. Figure 2 shows the PRISMA flow chart, which shows the step-by-step literature search and consideration/rejection procedure.

**Figure 2.**
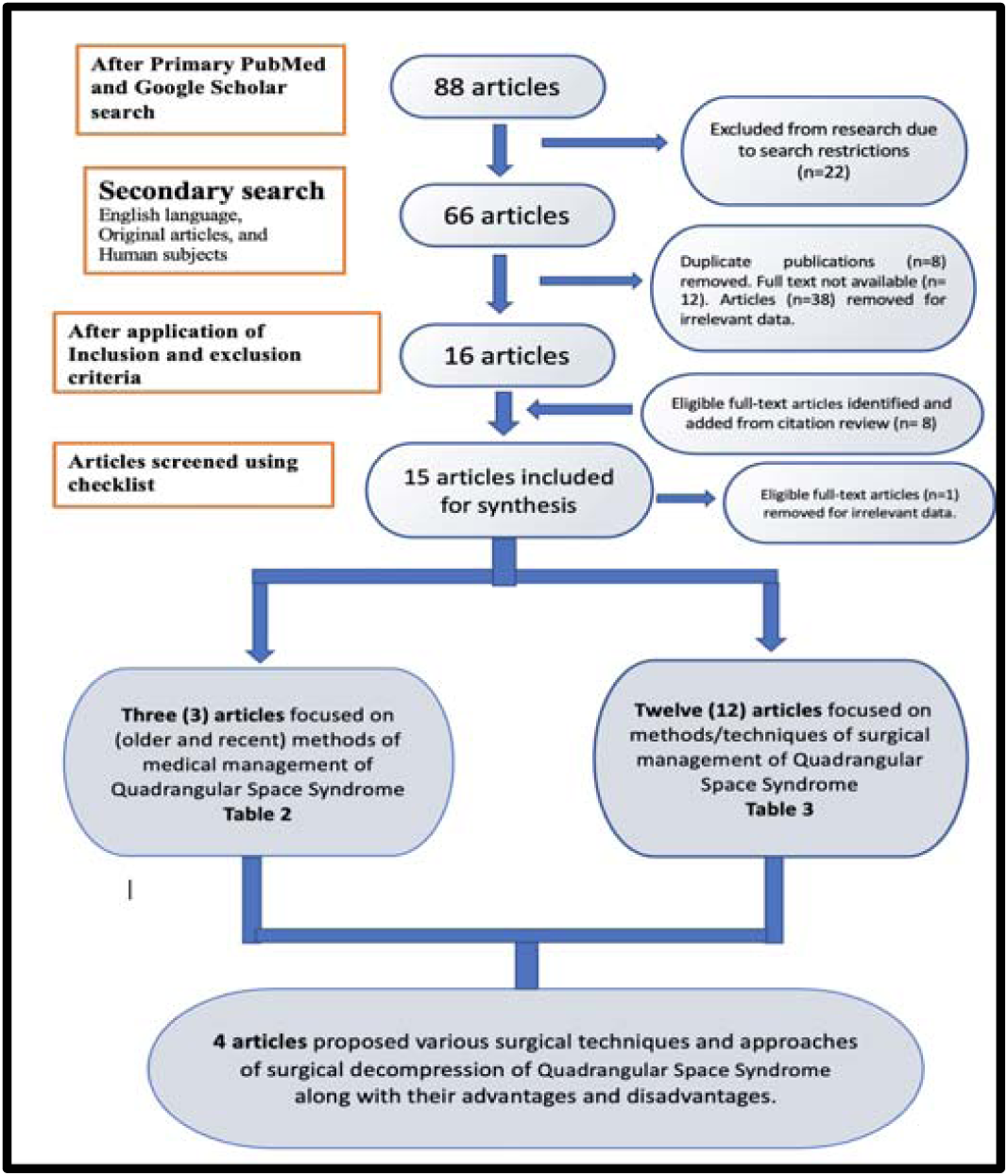
PRISMA flow chart of systematic literature search performed for publications focusing on methods of medical and surgical managemnt of Quadrangular Space Syndrome.

### 3.2 Characteristics of Included studies

**Table 2**, and **Table 3** summarizes the characteristics of the 15 selected studies. Out of the reviewed 15 publications, 3 focused on the methods (conservative and advanced) of medical management of QSS (Shown in Table 2). These 3 publications comprised of 2 case reports and 1 case study. Out of the selected 15 publications, 12 articles focused on methods/techniques (previous, recent, and newer) of surgical management of QSS (Shown in **Table 3**). These 12 publications comprised of 3 case series, 2 cadaveric study, 2 book chapters, 3 review articles, and 2 case reports.

**Table 2.**
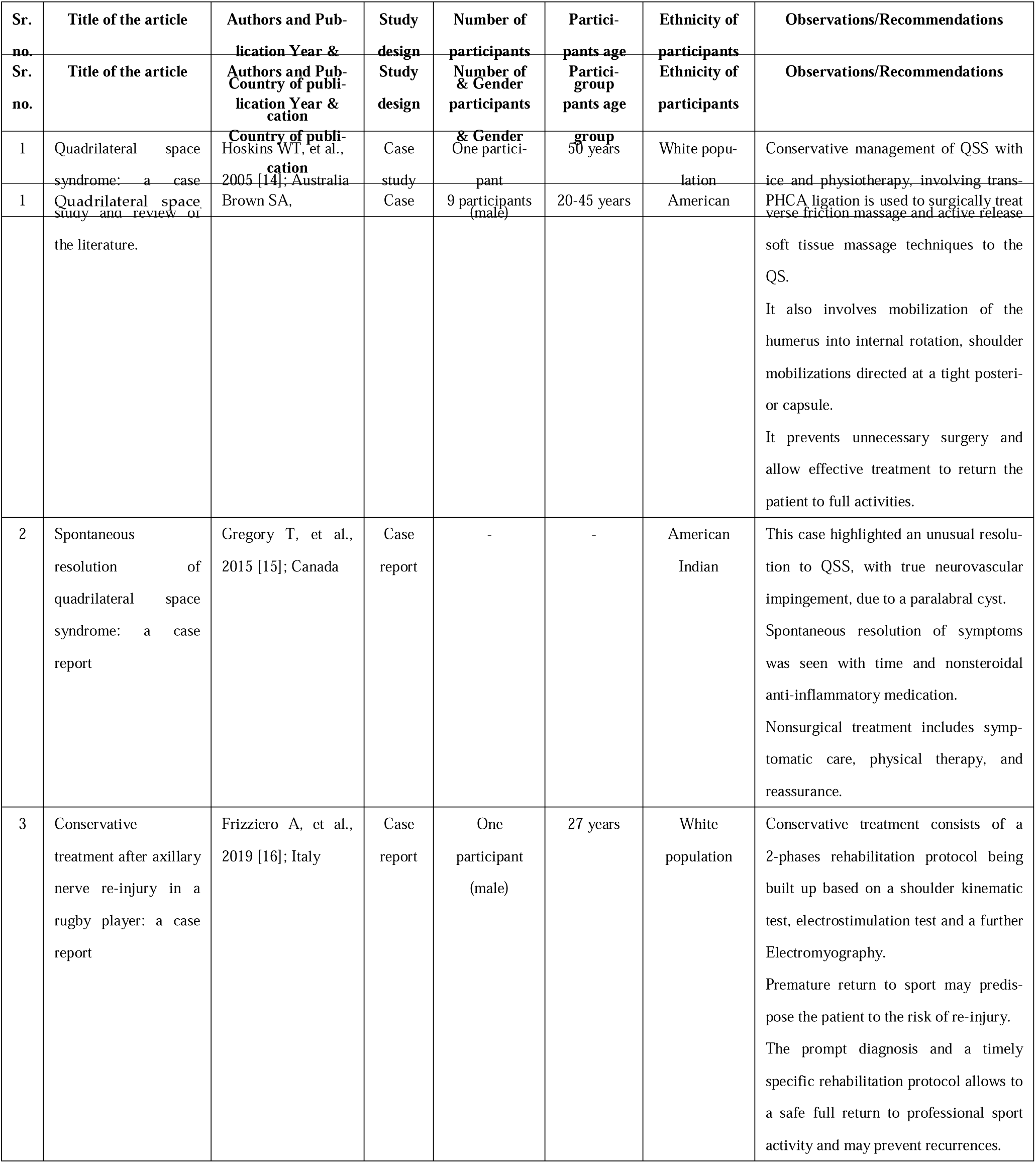
Characteristics of Three publications focusing on older and recent methods of medical management of Quadrangular Space Syndrome. QS: Quadrangular space; QSS: Quadrangular space syndrome

**Table 3.**
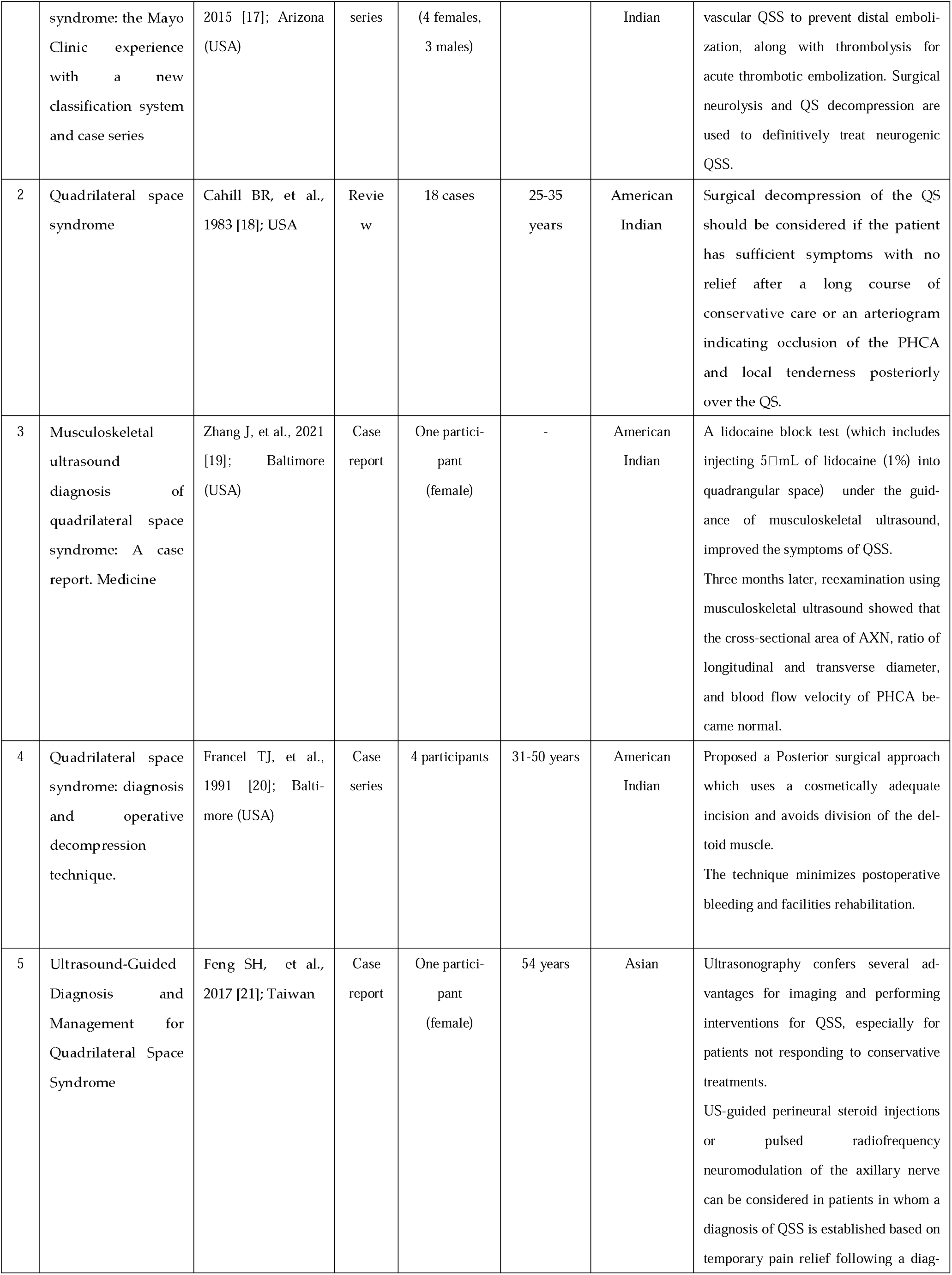
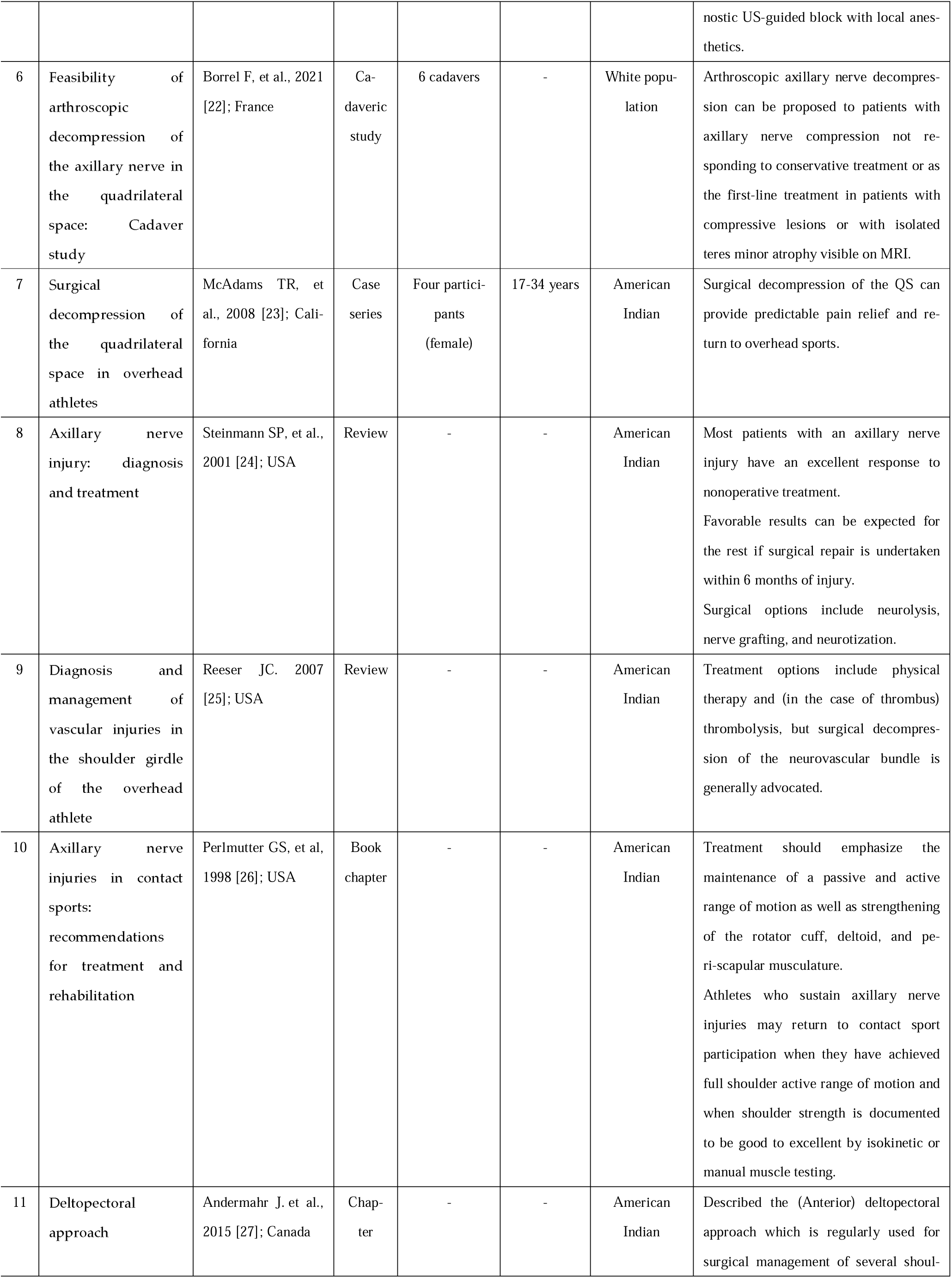
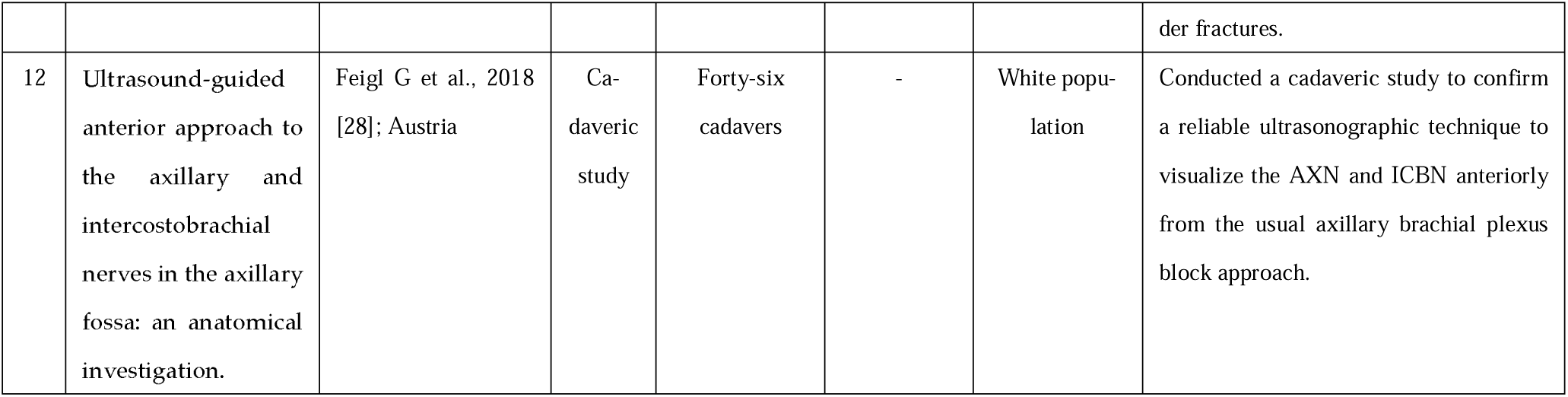
Characteristics of twelve publications focusing on methods/techniques of Surgical management of Quadrangular Space Syndrome. QS: Quadrangular space; QSS: Quadrangular space syndrome; AXN: Axillary nerve; PHCA: Posterior Circumflex Humeral Artery

Among the 15 reviewed articles, only 4 articles focused specifically on describing the various surgical techniques of surgical decompression of QSS along with their advantages and disadvantages. The step-by-step procedure of each proposed surgical approach is mentioned in the results section.

**Table 4** shows the comparison of the anterior and posterior surgical approaches described in these 4 articles. The parameters used for comparing these two surgical approaches were anatomical ease of identifying involved neuro-vascular structures, technical expertise required to execute the procedure, probabilities of injury to neuro-vascular structures, probabilities of postoperative fibrosis and other complications, and time duration required to complete the procedure.

**Table 4:**
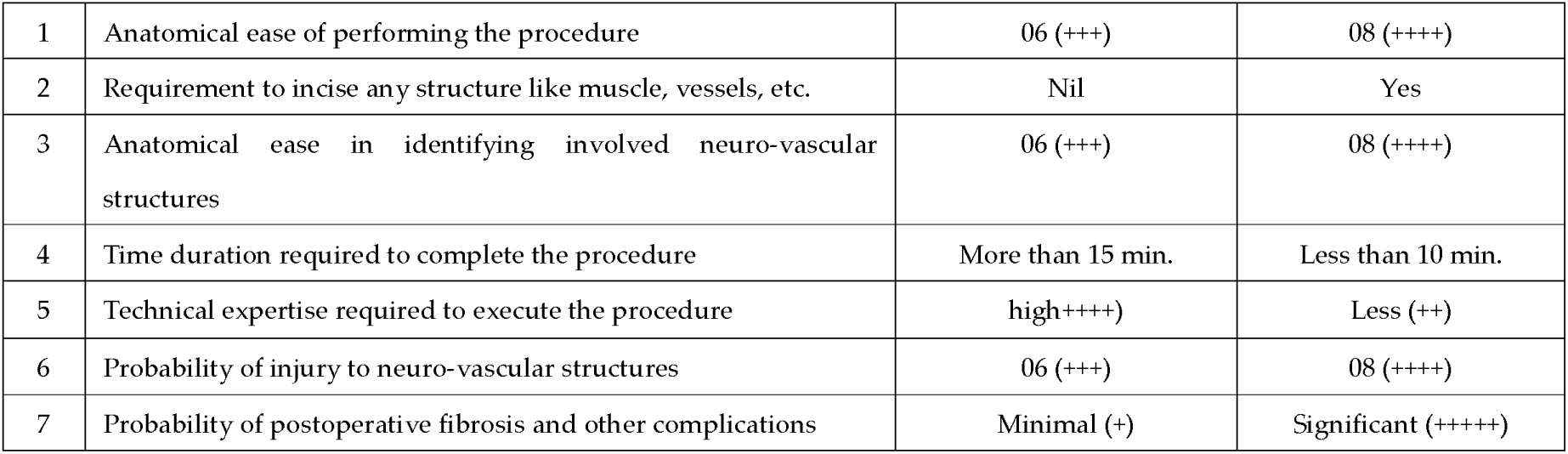
Comparison of both the approaches based on the studied parameters [29].

**Table 5:**
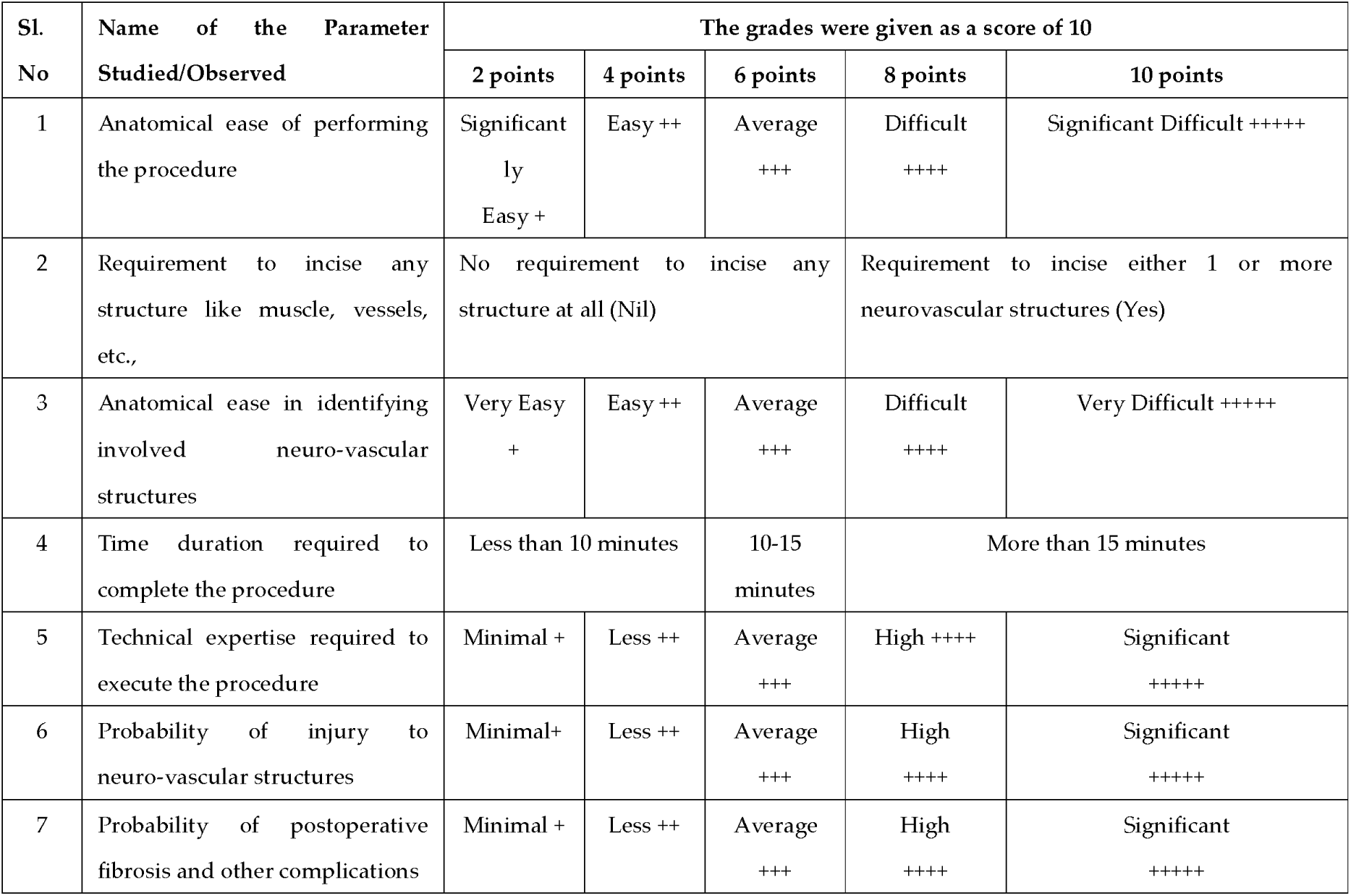
The grading system determined for every parameter studied is as follows [29].

### 3.3 Risk of Bias Assessment

The studies selected for this review study used a variety of methodological techniques. All three phases of the ROBIS tool were utilized to assess the risk of bias in our review study’s methodology. Except for a few points in Domain 2 of Phase 2, all other requirements were met. As a result, the total risk of bias was determined to be minimal.

### 3.4 Commonly used surgical approaches for surgical decompression in QSS are as follows

#### a. Anterior (Delto-pectoral) approach

For shoulder arthroplasty, the anterior approach is currently the preferred technique. The patient is positioned in a supine position with the right arm in 90-degree abduction in this method. A 12 cm long incision running from the lateral margin of the coracoid process to the proximal humeral shaft near to the deltoid tuberosity is to be taken on the right side, using bony and surface landmarks (acromion, clavicle, coracoid process, deltoid) [27]. Within the Delto-pectoral groove, the clavipectoral membrane is incised. The subscapularis muscle is exposed after retracting the deltoid muscle laterally and the conjoint tendon medially. The AXN can be found on the surface of the subscapularis muscle and close to its lower border. Because it is directly associated to the teres minor belly inferiorly, the lower border of the subscapularis muscle is the most critical region.

The lower border of the teres minor is detected, and quadrangular space is traced by inserting a fingertip horizontally forwards along the plane of the lower border of the teres minor, using rough dissection. The contents of the QS can be dissected using blunt dissection after the QS has been identified. The teres major muscle does not need to be exposed, and the contents of the QS (AXN and PHCA) can be recognized and traced within the QS via blunt dissection. Any fibrous strands or adhesions in the QS and its contents can be removed. This method spares no muscle [27].

#### b. Posterior/Scapular approach

The patient is put in a lateral decubitus position and a longitudinal incision of approximately 4 cm is made over the posterior shoulder in this surgical approach. The underlying fat within the QS between the teres and the teres major is shown by securing the posterior border of the deltoid and reflecting it supero-laterally. Following that, the AXN and posterior circumflex humeral vessels will be palpated as they exit the QS, and the QS will be located and secured [20].

## Discussion

In their work, **Cahill BR and Palmer RE** (1983) advocated a posterior technique in which a transverse incision was made parallel to the scapula’s spine and curved inferiorly over the rear portion of the humerus. The deltoid was later severed from the scapula’s spine. When Teres minor was inserted into the rotator cuff, it became separated and reflected medially. Blunt and sharp dissection were used to decompress quadrangular space. This method produced satisfactory results in 16 of 18 patients [18].

### Pitfalls of this technique

i. Removal of Deltoid and Teres Minor resulted in excess bleeding intra-operatively [18].
ii. Division of teres minor weakened the rotator cuff and the lateral arm rotation [18].
iii. The postoperative wide scar may itself compress the neurovascular bundle [18].
iv. Postoperative chronic pain and formation of poor quality tissue [18, 20]

**Francel TJ et al. (1991**) in their study used a vertical or S-shaped incision which was made on the region of highest tenderness, i.e., quadrangular space, and skin flaps were elevated to expose the inferior border of the deltoid, according to their study [20]. After incising the deltoid fascia and exposing the teres muscle bellies, the deltoid was retracted superiorly. The quadrilateral space was reached once the fascia between the teres muscle bellies was opened. This procedure did not separate the Deltoid and Teres Minor muscles. The AXN and PHCA were isolated and identified. The motor response of the teres minor and deltoid muscles was validated after nerve stimulation. Finger insertion divides fibrous bands and decompresses space [20].

### Advantages of this technique

i. Intact Deltoid and Teres Minor reduced bleeding and quick postoperative shoulder movement is possible [20].
ii. Fibrous atrophy of the deltoid was prevented [20].
iii. The postoperative scar was smaller [20].

In their study, Andermahr J. et al. (2015) emphasised the (anterior) deltopectoral technique, which was commonly employed for practically any shoulder fracture treatment and was frequently recommended, especially in anterior glenoid fractures [27]. This method, with a few tweaks, could be used to decompress the QSS surgically.

The (Anterior) Delto-pectoral technique was also employed by Feigl G et al. (2018) to view the AXN anteriorly. AXN was found near the inferolateral boundary of the subscapular muscle to enter the QS in 91 out of 92 limbs in their cadaveric investigation. The roof of the area is defined in this manner by the insertion of the subscapular muscle at the lesser tubercle [28].

## Limitations

A review of such items was impossible due to a lack of free access to whole text for various publications. Even abstracts were absent in a few papers. This constraint will be addressed in future review papers.

## 5. Conclusions

We conclude that the anterior approach is technically easier to perform and can be employed for surgical decompression in QSS based on our findings from our literature research. Furthermore, an ultrasound-guided anaesthetic block to AXN can be simply integrated with this method. The authors propose that cadaveric studies be used to give anatomists and surgeons more opportunities to perform and evaluate older and newer surgical techniques.

## Data Availability

All data produced in the present work are contained in the manuscript.

https://www.crd.york.ac.uk/prospero/.

## Author Contributions

The contributions from the authors of this research study are as follows: “Conceptualization, SC, SK, SM.; methodology, SC.; software, not applicable; validation, SC., SS. and SP; formal analysis, SC; investigation, SC; resources, SC, SK and SP.; data curation, SC and SP.; writing—original draft preparation, SC, SP, RJ, PA.; writing—review and editing, SC and SS.; visualization, SS.; supervision, SM.; project administration, SC, SK and SS.; funding acquisition, not applicable. All authors have read and agreed to the published version of the manuscript.”

## Institutional Review Board Statement

Since this was a systematic review which did not involve humans or animals, no ethical approval was required, and the AIIMS Rajkot Institutional Ethical Committee was not consulted. However, the AIIMS Rajkot Research Review Board was notified about this research work, along with the author information.

## Informed Consent Statement

Not applicable, as the study did not involve humans.

## Data Availability Statement

Not applicable.

## Acknowledgments

the authors wish to thank the faculties Dr. Pradip Chauhan, Assistant Professor, AIIMS Rajkot for his constant support whenever required in this review study.

## References

1. Miller RH, Azar FM, Throckmorton TW. Campbell’s Operative Orthopedics. Twelfth edition. Volume 3. Par Sports Medicine. Chapter 46. Shoulder and Elbow injuries. Page: 2238. https://www.medicalbook.co.in/medical-book-orthopedics.html.

2. Van de PD, Kuijer PP, Langenhorst T, Maas M. High prevalence of self-reported symptoms of digital ischemia in elite male volleyball players in the Netherlands: a cross-sectional national survey. Am J Sports Med. 2012;40(10):2296–302. https://doi.org/10.1177/0363546512456973.

3. Cormier PJ, Matalon TA, Wolin PM. Quadrilateral space syndrome: a rare cause of shoulder pain. Radiology. 1988;167(3):797–98. https://doi.org/10.1148/radiology.167.3.3363143.

4. McClelland D, Hoy G. A case of quadrilateral space syndrome with involvement of the long head of the triceps. Am J Sports Med. 2008;36(8):1615–17. https://doi.org/10.1177/0363546508321476.

5. Reutter D, Hunziker R, Husmann M. Computed angiogram of the upper extremities for diagnosing a rare cause of brachial arterial embolism: the “Pitcher Syndrome”. Eur Heart J. 2010;31(22):2782. https://doi.org/10.1093/eurheartj/ehq269.

6. Chautems RC, Glauser T, Waeber-Fey MC, Rostan O, Barraud GE. Quadrilateral space syndrome: case report and reviewof the literature. Ann Vasc Surg. 2000;14(6):673–76. https://doi.org/10.1007/s100169910120.

7. Snell Richard S. Clinical Anatomy by Regions. 9th edition. Chapter 9. The Upper Limb. Quadrangular space. 359. https://www.yumpu.com/en/document/view/55653009/snells-clinical-anatomy-by-regions-9th-edition.

8. Manske RC, Sumler A, Runge J. Quadrilateral space syndrome. Hum Kinet. 2009;14:45–47. https://dx.doi.org/10.1123%2Fatt.14.2.45.

9. Rea P. Essential clinically applied anatomy of the peripheral nervous system in the head and neck. Science Direct Neck. 3.15.1 Axillary Nerve. 2016. https://www.elsevier.com/books/essential-clinically-applied-anatomy-of-the-peripheral-nervous-system-in-the-head-and-neck/rea/978-0-12-803633-4.

10. Juneja P, Hubbard JB. StatPearls [Internet]. StatPearls Publishing; Treasure Island (FL): Aug 22, 2020. Anatomy, Shoulder and Upper Limb, Arm Teres Minor Muscle. https://www.ncbi.nlm.nih.gov/books/NBK513324/.

11. Elzanie A, Varacallo M. StatPearls [Internet]. StatPearls Publishing; Treasure Island (FL): Aug 22, 2020. Anatomy, Shoulder and Upper Limb, Deltoid Muscle. https://www.ncbi.nlm.nih.gov/books/NBK537056/

12. Whiting P, Savovic J, Higgins JP, et al. ROBIS: A new tool to assess risk of bias in systematic reviews was developed. J Clin Epidemiol. 2016;69:225–234. doi:10.1016/j.jclinepi.2015.06.005.

13. Dalagiannis N, Tranovich M, Ebraheim N. Teres minor and quadrilateral space syndrome: A review. J Orthop. 2020 Jan 21;20:144–146. doi: 10.1016/j.jor.2020.01.021.

14. Hoskins WT, Pollard HP, McDonald AJ. Quadrilateral space syndrome: a case study and review of the literature. Br J Sports Med. 2005 Feb;39(2):e9. doi: 10.1136/bjsm.2004.013367.

15. Gregory T, Sangha H, Bleakney R. Spontaneous resolution of quadrilateral space syndrome: a case report. Am J Phys Med Rehabil. 2015 Jan;94(1):e1–5. doi: 10.1097/PHM.0000000000000237.

16. Frizziero A, Vittadini F, Del Felice A, Creta D, Ferlito E, Gasparotti R, Masiero S. Conservative treatment after axillary nerve re-injury in a rugby player: a case report. Eur J Phys Rehabil Med. 2019 Aug;55(4):510–514. doi: 10.23736/S1973-9087.18.05165-1. Epub 2018 Dec 21.

17. Brown SA, Doolittle DA, Bohanon CJ, Jayaraj A, Naidu SG, Huettl EA, Renfree KJ, Oderich GS, Bjarnason H, Gloviczki P, Wysokinski WE, McPhail IR. Quadrilateral space syndrome: the Mayo Clinic experience with a new classification system and case series. Mayo Clin Proc. 2015 Mar;90(3):382–94. doi: 10.1016/j.mayocp.2014.12.012. Epub 2015 Jan 31.

18. Cahill BR, Palmer RE. Quadrangular space syndrome. J Hand Surg Am. 1983;8(1):65–69. https://doi.org/10.1016/s0363-5023(83)80056-2.

19. Zhang J, Zhang T, Wang R, Wang T. Musculoskeletal ultrasound diagnosis of quadrilateral space syndrome: A case report. Medicine (Baltimore). 2021;100(10):e24976. doi:10.1097/MD.0000000000024976

20. Francel, Thomas J. M.D.; Dellon, A Lee M.D.; Campbell, James N. M.D. Quadrilateral Space Syndrome: Diagnosis and Operative Decompression Technique, Plastic and Reconstructive Surgery: May 1991 - Volume 87 - Issue 5 - p 911–916. https://doi.org/10.1097/00006534-199105000-00016.

21. Feng SH, Hsiao MY, Wu CH, Özçakar L. Ultrasound-Guided Diagnosis and Management for Quadrilateral Space Syndrome. Pain Med. 2017 Jan 1;18(1):184–186. doi: 10.1093/pm/pnw256.

22. Borrel F, Desmoineaux P, Delcourt T, Pujol N. Feasibility of arthroscopic decompression of the axillary nerve in the quadrilateral space: Cadaver study. Orthop Traumatol Surg Res. 2021 Feb;107(1):102762. doi: 10.1016/j.otsr.2020.102762. Epub 2020 Dec 14.

23. McAdams TR, Dillingham MF. Surgical decompression of the quadrilateral space in overhead athletes. Am J Sports Med. 2008 Mar;36(3):528–32. doi: 10.1177/0363546507309675. Epub 2007 Nov 30.

24. Steinmann SP, Moran EA. Axillary nerve injury: diagnosis and treatment. J Am Acad Orthop Surg. 2001 Sep-Oct;9(5):328–35. doi: 10.5435/00124635-200109000-00006.

25. Reeser JC. Diagnosis and management of vascular injuries in the shoulder girdle of the overhead athlete. Curr Sports Med Rep. 2007 Oct;6(5):322–7. PMID: 17883968

26. Perlmutter GS, Apruzzese W. Axillary nerve injuries in contact sports: recommendations for treatment and rehabilitation. Sports Med. 1998 Nov;26(5):351–61. doi: 10.2165/00007256-199826050-00005.

27. Andermah Jr, McKee M, Nam D. AO Surgery Reference. First edition. 2015. Deltopectoral approach. https://surgeryreference.aofoundation.org/orthopaedic-trauma/adulttrauma/clavicle/additionalcredits. https://surgeryreference.aofoundation.org/orthopedic-trauma/adult-trauma/scapula/approach/deltopectoral-approach.

28. Feigl G, Aichner E, Mattersberger C, Zahn PK, Avila Gonzalez C, Litz R. Ultrasound-guided anterior approach to the axillary and intercostobrachial nerves in the axillary fossa: an anatomical investigation. Br J Anaesth. 2018;121(4):883–89. https://doi.org/10.1016/j.bja.2018.06.006.

29. Charmode S, Mehra S, Kushwaha S. REVISITING the Surgical Approaches to Decompression in Quadrilateral Space Syndrome: A Cadaveric Study. Cureus. 2022 Feb 26;14(2):e22619. doi: 10.7759/cureus.22619.

